# Facilitators and Barriers to Compliance with COVID-19 Guidelines: A Structural Topic Modelling Analysis of Free-Text Data from 17,500 UK Adults

**DOI:** 10.1101/2021.06.28.21259621

**Authors:** Liam Wright, Elise Paul, Andrew Steptoe, Daisy Fancourt

**Author notes:** Corresponding author: Liam Wright, 1-19 Torrington Place, London, WC1E 7HB.

## Abstract

**Background:** During the COVID-19 pandemic, the UK government has implemented a series of guidelines, rules, and restrictions to change citizens’ behaviour to tackle the spread of the virus, such as the promotion of face-masks and the imposition of lockdown stay-at-home orders. The success of these measures requires active co-operation on the part of citizens, but compliance has not been complete. Detailed data is required on the factors aiding or hindering compliance with these measures.

**Methods:** To understand the facilitators and barriers to compliance with COVID-19 guidelines, we used structural topic modelling, a text mining technique, to extract themes from over 26,000 free-text survey responses from 17,500 UK adults, collected between 17 November and 23 December 2020.

**Results:** The main factors facilitating compliance were desires to reduce risk to one’s self and one’s family and friends and to, a lesser extent, the general public. Also of importance were a desire to return to normality, the availability of activities and technological means to contact family and friends, and the ability to work from home. Identified barriers were difficulties maintaining social distancing in public (due to the actions of other people or environmental constraints), the need to provide or receive support from family and friends, social isolation, missing loved one, and mental health impacts, perceiving the risks as low, social pressure to not comply, and difficulties understanding and keep abreast of changing rules. Several of the barriers and facilitators raised were related to participant characteristics. Notably, women were more likely to discuss needing to provide or receive mental health support from friends and family.

**Conclusion:** The results demonstrate an array of factors contribute to compliance with guidelines. Of particular policy importance, the results suggest that government communications that emphasizes the potential risks of COVID-19 and provides simple, consistent guidance on how to reduce the spread of the virus would improve compliance with preventive behaviours.

## Background

To tackle the COVID-19 pandemic, governments have focused on reducing transmission of the virus by influencing citizens’ behaviour. Governments have mandated the wearing of face masks in public places, recommended social distancing, promoted regular hand-washing, and even ordered the closing of businesses, prohibited household mixing, and implemented stay-at-home “lockdown” orders. Where followed, these measures are effective at reducing infection rates [1]. But while compliance has been high overall, it has not been complete [2, 3], and levels of compliance have in general decreased since the start of the pandemic [4, 5]. Promoting compliance with preventive behaviours has been an important component of efforts to tackle the pandemic. For example, in the UK, some measures have been backed with the force of law, with non-compliers subjects to fines or criminal sentences. Public health messaging has also been widely used, emphasising the need to save lives and protect the National Health Service (NHS).

Policymakers’ beliefs about compliance behaviour – the extent to which citizens comply, for how long, and in which contexts – have influenced the measures that have been put in place. For instance, the possibility of “risk compensation” [6] – individuals offsetting one risk-reducing behaviour with riskier behaviours elsewhere – was central to debates on making face masks compulsory in public places [7]. More controversially, the possibility of (behavioural) “fatigue” was cited as a reason to delay lockdown in the UK, fearing that citizens would not sustain compliance over a sufficiently long period of time [8]. Although this argument was widely criticised by behavioural scientists as lacking clarity and scientific support [9–11], it has received limited direct empirical testing to date [4, 5, 12].

Given the importance of citizens’ behaviour for tackling the current – and previous – pandemics, a large literature has grown on the determinants of compliance, including on the motivations, barriers, and facilitators of compliance and the personal and situational characteristics associated with high compliance levels [for reviews, see 13–15]. Recent work has shown a role of gender [14], worries about the virus [16, 17], pro-social motivations [16, 18], attitudes to risk [19], and assorted personality traits [20, 21] in predicting compliance during COVID-19. Several theoretical frameworks have been used to understand individual differences in compliance with preventive behaviours. One fruitful framework has been the COM-B model [16, 22]. The COM-B model [23, 24] posits that behaviour results from the interaction of physical and psychological attributes of the individual (Capability), autonomic and reflexive mental processes directing and energising behaviour (Motivation), and physical and social attributes of the environment (Opportunity). For instance, a person may enact a specific preventive behaviour (e.g., social distancing) if they have the knowledge that the behaviour is effective (psychological capability), are worried about catching COVID-19 (reflexive motivation), and do not perceive a strong social norm to act otherwise (social opportunity).

An issue with the literature on the determinants of compliance is that most studies use quantitative data. A limitation of this is that the factors studied are restricted to those the researcher has thought of in advance. Moreover, in many of these studies, the specific reasons why the studied factors are related to compliance has not been empirically examined. For instance, several studies show a link between confidence or trust in government and compliance behaviour [25–27], but the specific barriers or motivations to compliance that may be influenced by trust are not explored. This limits the lessons that can be drawn.

Qualitative studies offer an opportunity to deepen our understanding of compliance behaviour, allowing for greater flexibility in the exploration and identification of relevant phenomena. Recent qualitative studies from the UK find evidence of “alert fatigue”, with individuals expressing difficultly keeping abreast of frequently changing guidelines and, as a result, inadvertently breaking (or knowingly bending) government rules [28, 29]. It is difficult to imagine a purely quantitative study that would have identified these phenomena. Nevertheless, the small sample sizes typical of qualitative studies also limit the questions that can be asked – notably, those that statistically relate participants’ characteristics to the themes they discuss.

Text-mining approaches offer a middle ground, allowing for the extraction of themes from large-scale free-text data that can then be related to document metadata, such as the date the text was written and the characteristics of its author (e.g., their age, sex, or personality traits). Participants can provide spontaneous information, which can be summarized quantitatively and analysed as any other quantitative variable (e.g., in a regression model). In this study, we used structural topic modelling (STM) [30] – a text-mining technique – to identify barriers to compliance from free-text responses from over 17,500 UK adults during the COVID-19 pandemic, relating these to participants’ demographic, socioeconomic and personality characteristics identified as predictors of compliance behaviour in the wider compliance literature.

## Methods

### Participants

We used data from the COVID-19 Social Study; a large panel study of the psychological and social experiences of over 70,000 adults (aged 18+) in the UK during the COVID-19 pandemic. The study commenced on 21 March 2020 and involved online weekly data collection for 22 weeks with monthly data collection thereafter. Data collection is still ongoing. The study is not random and therefore is not representative of the UK population, but it does contain a heterogeneous sample. The sample was recruited using three primary approaches. First, convenience sampling was used, including promoting the study through existing networks and mailing lists (including large databases of adults who had previously consented to be involved in health research across the UK), print and digital media coverage, and social media. Second, more targeted recruitment was undertaken focusing on (i) individuals from a low-income background, (ii) individuals with no or few educational qualifications, and (iii) individuals who were unemployed. Third, the study was promoted via partnerships with third sector organisations to vulnerable groups, including adults with pre-existing mental health conditions, older adults, carers, and people experiencing domestic violence or abuse. Full details on sampling, recruitment, data collection, data cleaning and sample demographics are available at https://github.com/UCL-BSH/CSSUserGuide. The study was approved by the UCL Research Ethics Committee [12467/005] and all participants gave informed consent.

A one-off module on compliance behaviour was included in the survey between 17 November and 23 December 2020. The module included two free-text questions on facilitators and barriers to compliance, respectively:

1. Since the pandemic started, what has been encouraging or helping you to follow the guidelines, even if only to a partial extent?
2. If you have not been entirely following the guidelines, what are the factors that have been hindering you or acting as obstacles?

We conducted analyses for both questions, separately. 25,051 individuals participated in the data collection containing the free-text survey module (34% of participants with data collection by 23 December 2020). Responses to the free-text questions were optional. 18,742 participants provided a response to the question on facilitators of compliance (74.82% of eligible participants) and 11,902 participants recorded a response to the question on barriers (47.51% of eligible participants). Of these, 16,512 (88.1%) and 9,720 (81.6%) participants provided a valid record, the definition of which is provided below.

The period 17 November – 23 December 2020 overlaps with the beginning of the second wave in the UK. Government rules changed across the study period. A description of the changes in these rules is provided in the Supplementary Information.

### Data cleaning

We performed topic modelling using unigrams (single words). Responses were cleaned using an iterative process. We excluded responses containing fewer than five words and removed words that appeared in fewer than five responses. We also removed common “stop” words (“the”, “and”, “I”, etc.) from the analysis. We used complete case data so excluded a small number of participants with missing data on any covariate used in the analysis. Spelling mistakes were identified with the hunspell algorithm [31], amended manually if they had five or more occurrences, and replaced using the hunspell suggested word function if the number of occurrences was fewer than five. Where the algorithm provided multiple suggestions, the word with the highest frequency in the original dataset was used. We concatenated frequently-used multi-word concepts into single phrases where we deemed this to be important to our research question (e.g., “high risk”, “pre pandemic”). To reduce data sparsity, in the structural topic models, we used word stemming using the Porter [32] algorithm. Data cleaning was carried out in R version 3.6.3 [33].

### Data analysis

We performed several quantitative analyses. First, as not all participants chose to provide a response, we ran a logistic regression model to explore the predictors of providing a free-text response. Second, we used STM, implemented with the stm R package [34], to extract topics from responses, with the analysis carried for each question separately. STM treats documents as a probabilistic mixture of topics and topics as a probabilistic mixture of words. It is a “bag of words” approach that uses correlations between word frequencies within documents to define topics. STM allows for inclusion of covariates in the estimation model, such that the estimated proportion of a text devoted to a topic can differ according to document metadata (e.g., characteristics of its author). We included participant’s gender, ethnicity (white, non-white), age (basis splines, degrees of freedom 4), education level (degree or above, A-Level or equivalent, GCSE or below), living arrangement (alone; alone, without child; alone, with child), psychiatric diagnosis (any vs. none), long-term physical health conditions (0, 1, 2+), self-isolation status (yes vs no), and Big-5 personality traits (Openness to Experience, Conscientiousness, Extraversion, Agreeableness, Neuroticism; BFI-2 [35]), each collected at first data collection, and confidence in government to handle the pandemic (1 “None at all” – 7 “Lots”) collected during the same data collection as the free-text responses. For confidence in government, participants from devolved nations were asked about their home government, while those from England were asked about the central UK government. There was only a small amount of item missingness, so we used complete case data. More detail on the variables used in this analysis is provided in the Supplementary Information.

We ran STM models from 2-30 topics for each question and selected the final models based on visual inspection of the semantic coherence and exclusivity of the topics and close reading of exemplar documents representative of each topic. Semantic coherence measures the degree to which high probability words within a topic co-occur, while exclusivity measures the extent to that a topic’s high probability words have low probability for other topics. After selecting a final model, we carried out two further analyses. First, we decided upon narrative descriptions for the topics based on high probability words, high “FREX” words (a weighted measure of word frequency and exclusivity), and exemplar texts (responses with a higher proportion of text estimated for a given topic). Second, we ran regression models estimating whether topic proportions were related to author characteristics defined above. Third, we ran a regression model estimating whether self-reported adherence to COVID-19 guidelines (Are you following the recommendations from authorities to prevent spread of Covid-19? 1 “Not at all” - 7 “Very much so”) was related to topic proportions, to explore which barriers to compliance may make compliance particularly challenging.

Data cleaning and analysis was carried out by LW. LW and EP selected the number of model topics and LW, EP, AS and DF agreed upon narrative titles for the topics. Due to stipulations set out by the ethics committee, the free-text data cannot be made publicly available. The code used in the analysis is available at https://osf.io/nf4m9/.

### Role of the Funding Source

The funders had no final role in the study design; in the collection, analysis, and interpretation of data; in the writing of the report; or in the decision to submit the paper for publication. All researchers listed as authors are independent from the funders and all final decisions about the research were taken by the investigators and were unrestricted.

## Results

### Descriptive Statistics

16,512 participants provided a valid free-text response to the question on facilitators, and 9,720 individuals provided a valid free-text response to the question on barriers (17,706 unique individuals overall). Descriptive statistics for respondents are displayed in Table 1, with figures for the total sample also shown for comparison. Participants in the COVID-19 Social Study are disproportionately female, older age, and more highly educated, relative to the general population [36]. There were some differences between those who provided a (valid) response and those that did not. Figure S1 displays the results of logistic regression models exploring the predictors of providing a response. Responders to the question on facilitators had higher than average levels of compliance with COVID-19 guidelines, while responders to the question on barriers had lower than average compliance levels. Responders for both questions were disproportionately female, more highly educated and had lower confidence in government, on average. There were also differences according to health and personality traits, though the direction of the association differed by question.

**Table 1:**
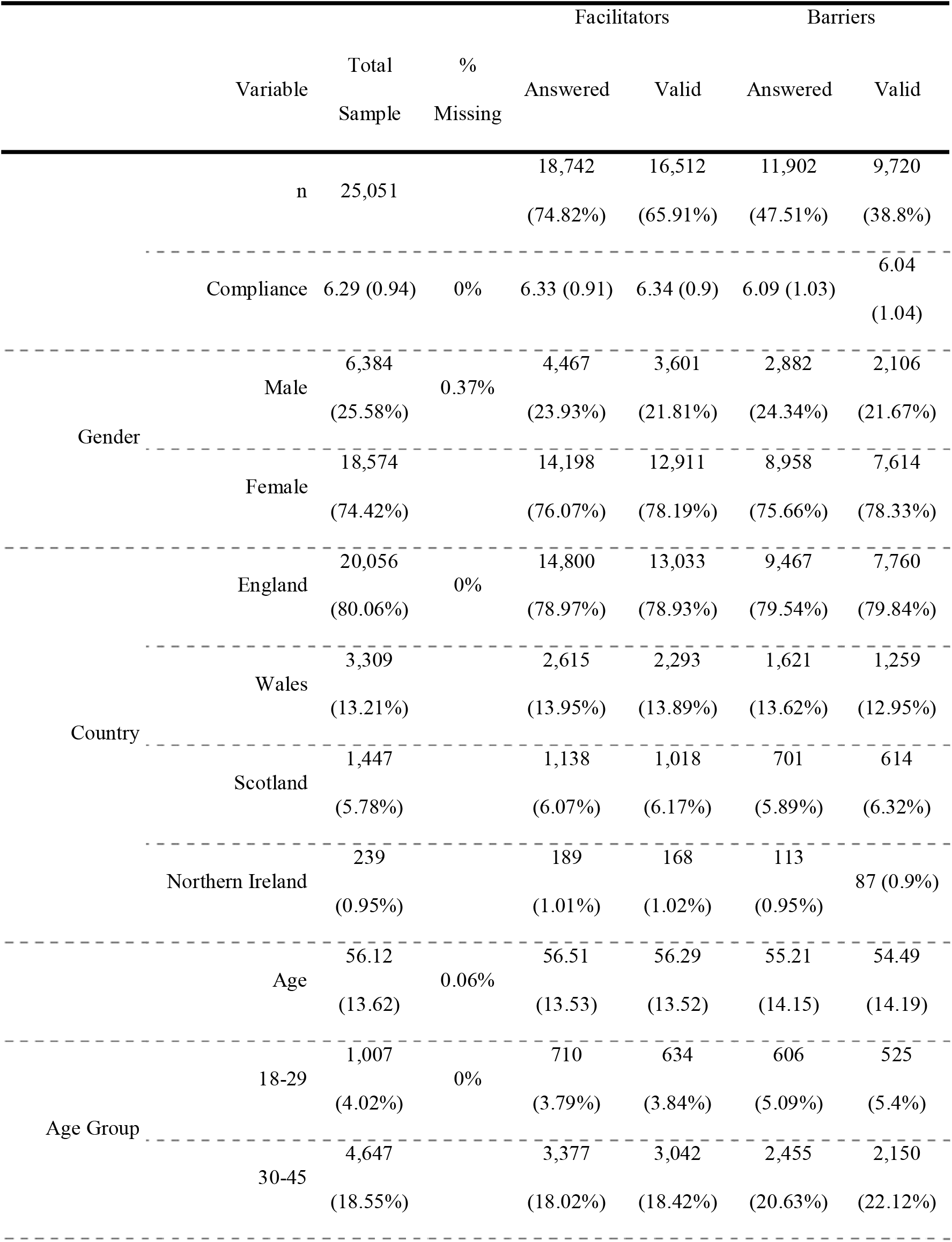
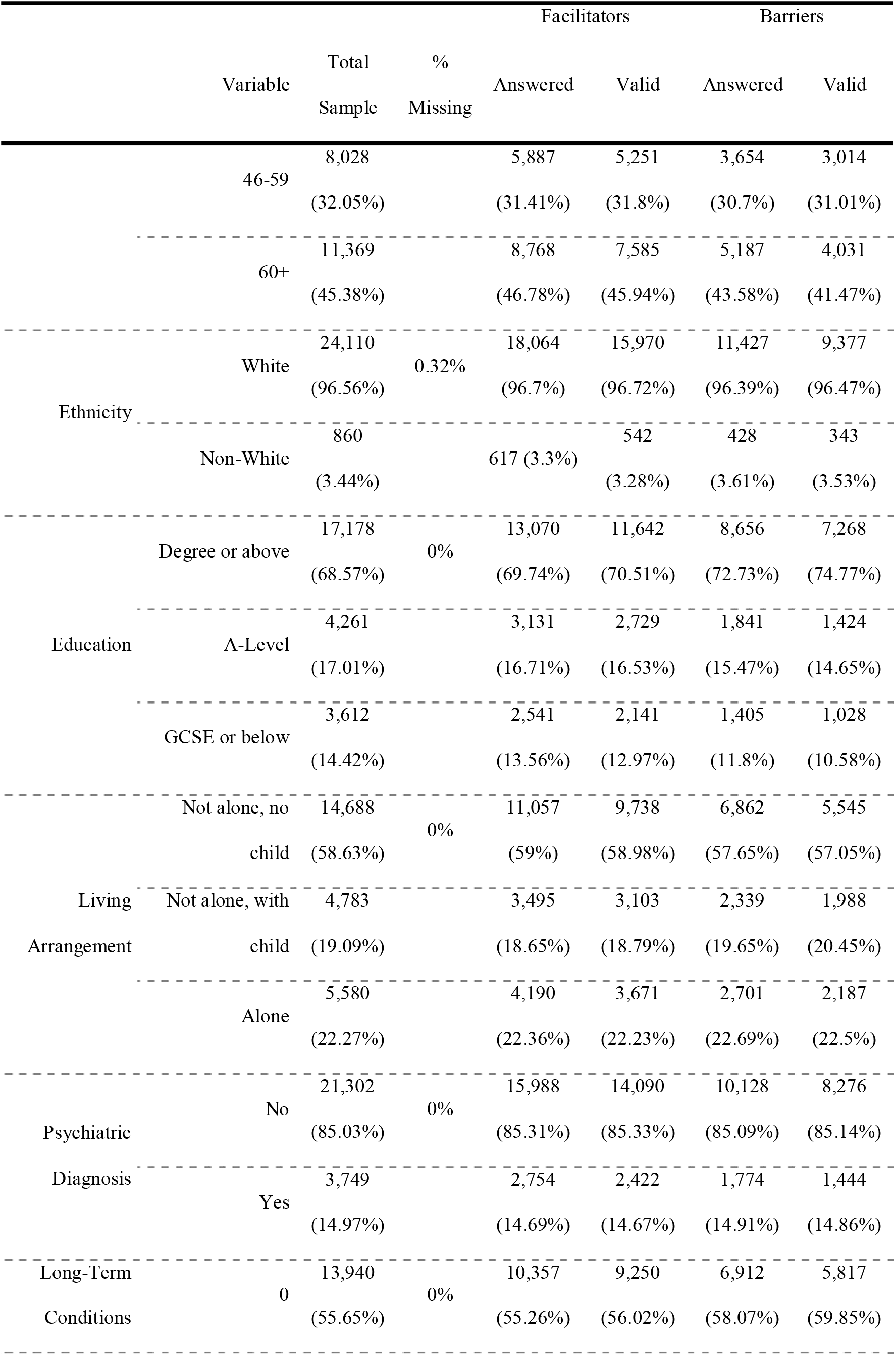
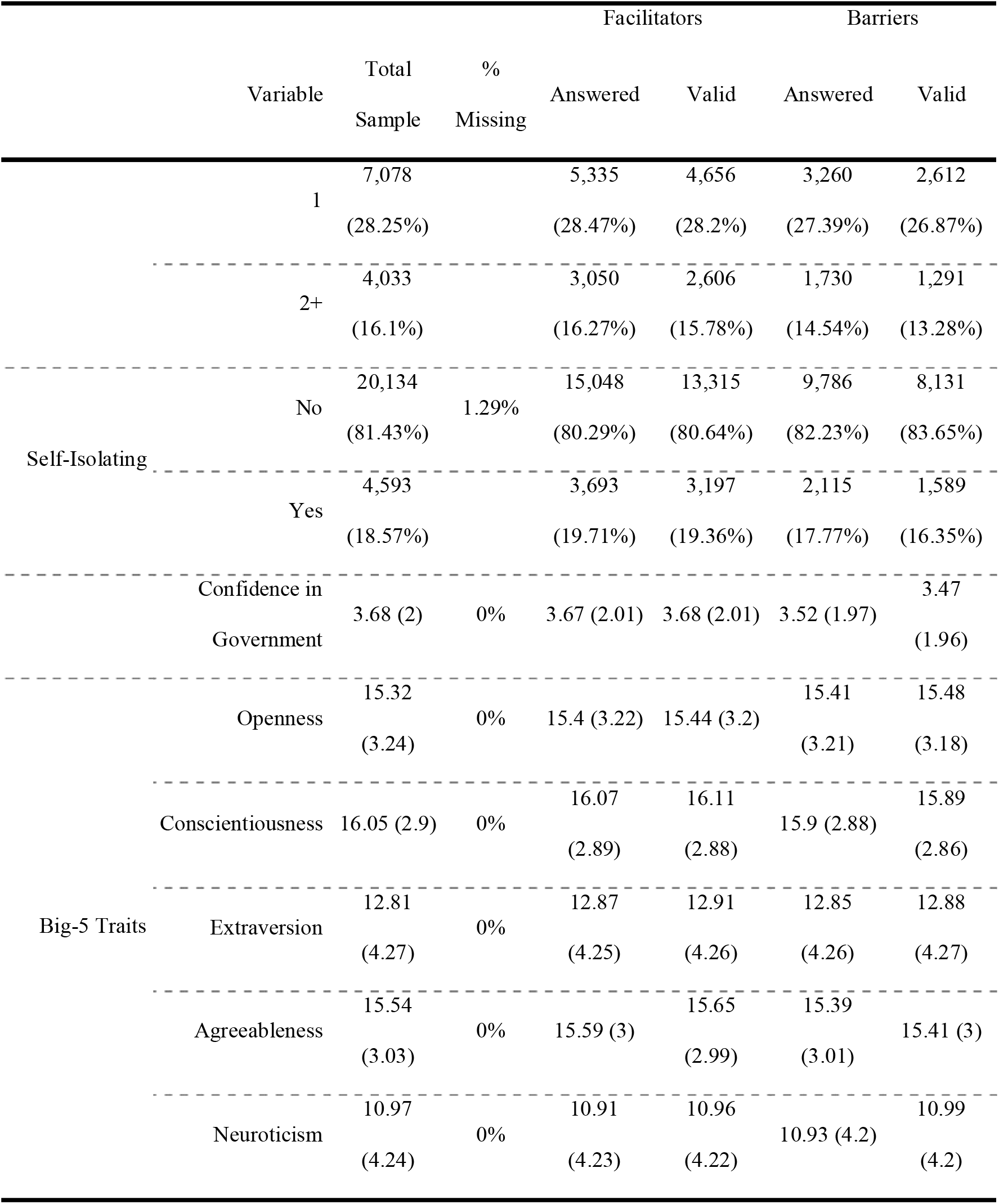
Descriptive Statistics

A word cloud of the twenty most frequently used words for each question is displayed in Figure 1. For facilitators, the most used words typically related to worries about catching the virus and protecting family and friends. For barriers, the most used words related to social distancing, mental health, and family and friends.

**Figure 1:**
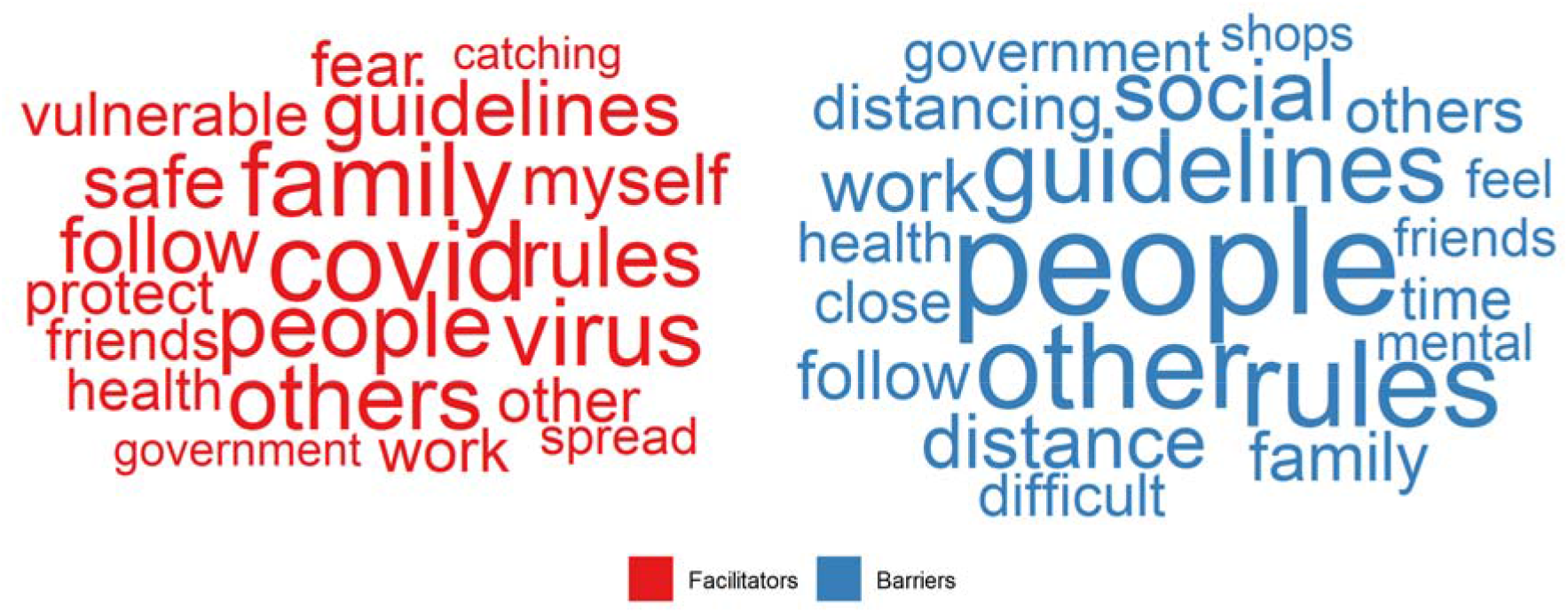
Word cloud. Twenty most frequently used words across responses, by question. Words sized according to proportion of responses they appear in.

### Facilitators of Compliance

A 14-topic solution was chosen to categorise facilitators of complying with guidelines. Exemplar quotes, topic descriptions, and topic proportions are presented in Table 2. Topics are ordered according to the estimated proportion of text devoted to each topic. There was some overlap in these topics in the themes they identified and there were also some cases of topics containing multiple themes.

**Table 2:** Topic descriptions – facilitators of compliance

| Topic | Proportion | Short Title | Description | FREX | Exemplar Text |
| --- | --- | --- | --- | --- | --- |
| F1 | 12.17% | Catching and transmitting COVID | Fear of catching COVID and infecting others, especially parents | fear, pass, catch, elderli, rel, dy, myself, covid, contract, longcovid | “I am terrified of being a vector for the illness and very scared of catching it myself. I particularly fear passing it on to my parents and inlaws who are all in their 70a [sic]” |
| F2 | 10.04% | Protecting high risk | Worry about catching COVID as high risk or relatives high risk | viru, underli, risk, spread, prevent, potenti, minimis, highrisk, reduc, condit | “fear of catching the virus myself or transmitting the virus eg to my 92 yo father or my partner with multiple underlying health conditions.” |
| F3 | 9.33% | Public information | Consuming media on COVID, such as government briefings | new, advic, inform, listen, scientist, tv, media, scientif, trust, watch | “The news on the T.V. The news on the Internet. Sometimes the governments bulletins. Always Listening and reading what Sir Patrick Vallance. Prof Chris Whitty. Prof Van Tam. have to say.” |
| F4 | 8.39% | Following the rules | Rule following as a personality trait. Also contains text on instrumental rule following | follow, rule, guidelin, stick, peopl, make, upset, consider, reason, adher | “I tend to just stick to rules even if I don't necessarily agree with them”<br>“for me, rules are rules....I may not agree with them or believe they have the outcome they suggest, but it's the rules so I follow along” |
| F5 | 7.62% | Social responsibility | Acting out of common sense and sense of duty and civic responsibility | common, sens, respons, infect, duti, commun, civic, other, act, respect | “Common sense and a sense of responsibility to others regardless of their attitude [sic]” |
| F6 | 6.96% | Protecting vulnerable | Reducing risks for those with existing clinical vulnerabilities | vulner, extrem, clinic, protect, categori, father, husband, older, chronic, mum | “To protect myself (clinically vulnerable), my husband (same), and 3 parents (clinically extremely vulnerable).” |
| F7 | 6.62% | Reminders and accessibility | Availability of hand sanitizer and reminders to wear masks | keep, mask, wear, hand, wash, sanitis, facemask, safe, transport, sign | “Signs on shop/building entrances reminding to wear mask, provision of hand sanitiser” |
| F8 | 6.49% | Safety of loved ones | Protecting family and friends | famili, safeti, concern, healthi, ill, encourag, friend, self, safeguard, fall | “To keep my family safe. I couldn't bear to be responsible for someone in my family falling ill with COVID-19.” |
| F9 | 6.37% | Return to normality | Following guidelines to end pandemic sooner and return society to normal. | normal, vaccin, hope, sooner, life, quickli, quicker, control, end, desir | “The hope of the pandemic ending and life being able to return to more of a normality” |
| F10 | 5.76% | Working from home / support bubbles | Ability to stay at home due to unemployment or working arrangement. Benefit of having support bubbles. | work, carehom, babi, bubbl, home, employ, ward, afford, alon, support | “Being in a support bubble as I live alone. If we did not have support bubbles I think I would have had to break the restrictions for my own mental wellbeing.”<br>“I prefer to be at home and because I can work from home I have not had to go out much.” |
| F11 | 5.5% | Activities and Zoom | Availability of activities, including walking and going out in nature. Speaking to family, friends and online groups over Zoom. | garden, meet, walk, exercis, weather, easier, outdoor, zoom, enjoi, dog | “a structured exercise routine i.e. online Zoom classes, weekly exercise classes outside in local park and regular long cycle rides and walks” |
| F12 | 5.48% | COVID symptoms | Discussion of COVID symptoms and test results | test, april, month, march, result, surgeri, diagnos, posit, week, flu | “I was told to isolate for 6 days due to test and trace app” |
| F13 | 5.1% | Protecting the NHS | Reducing burden on NHS and its workers | nh, impact, pressur, know, worker, servic, burden, motiv, understand, frontlin | “Husband frontline NHS worker. I have heard how bad it can be for patients and staff”<br>“Knowing that I would be helping the NHS to not become overwhelmed with patients” |
| F14 | 4.16% | Miscellaneous themes | Gallimaufry of themes, several related to those identified in Topics 1-13. | children, school, teacher, kid, young, adult, appli, grandchildren, socialis, differ | - |

Several of the topics related to desires to protect oneself and others from COVID-19. The largest topic, Topic F1 (12.17%; Catching and transmitting COVID), included text on worries about catching the virus and passing it on to family members, particularly elderly parents. Topics F2 (10.04%; Protecting high risk) and Topic F6 (6.96%; Protecting vulnerable) related to protecting high risk and clinically vulnerable individuals, specifically. Exemplar texts typically referred to reducing risk for oneself and for one’s family and to a lesser extent the wider population. Similar to Topic F1, Topic F8 (6.49%; Safety of loved ones) related to protecting family and friends (topics differ in specific words used). Topic F13 (5.1%; Protecting the NHS) related to reducing the burden on the National Health Service and its workers.

Prosocial motivations were also identified in Topic F5 (7.62%; Social responsibility), which explicitly couched compliance as a matter of social responsibility, civic duty or simply a matter of “common sense”. Topic F4 (8.39%; Following the rules) identified individuals predisposed to follow rules in general, though this topic also surfaced responses from individuals describing others’ rule following as a motivator. Topic F9 (6.37%; Return to normality) related to desires to hasten the end of the pandemic and return to life as normal.

A role of media coverage and scientific information was identified in Topic F3 (9.33%; Public information), which contained text on news and media reports, including briefings from Government ministers and scientists. Topics F7, F10 and F11 each related to factors that made compliance easier. Topic F7 (6.62%; Reminders and accessibility) included positive statements on the availability of hand sanitiser in shops and on reminders to wash hands and wear masks in public places. Topic F10 (5.76%; Working from home/support bubbles) included responses from individuals who had been able to work from home (or who had lost work and so did not need to travel to a workplace) and also on the availability of support bubbles, allowing participants to receive or give support to family and friends. Topic F11 (5.5%; Activities and Zoom) related to activities that had improved quality of life during lockdown, including walking in nature and participating in online activities, such as arts and talking to family members over Zoom.

The final two topics, Topics F12 and F14, contained irrelevant material or identified texts on a heterogeneous set of themes. Topic F12 (5.48%; COVID symptoms) included discussion of personal symptoms from COVID-19 or experiences with the test and trace system. Responses appeared to arise from participants expanding on their response to preceding questionnaire items. Topic F14 (4.16%; Miscellaneous themes) identified responses generally covering themes from the other topics, typically using slightly different phrasing.

### Compliance Facilitators and Respondent Characteristics

Figures 2-4 display the results of models regressing topic proportions on respondent characteristics. Figure 2 shows associations with age. Figure 3 shows associations with personality traits. Figure 4 shows associations with demographics, socio-economic factors, health and participants’ confidence in government. (Note, each of these results come from models with adjustment for other measured factors.)

**Figure 2:**
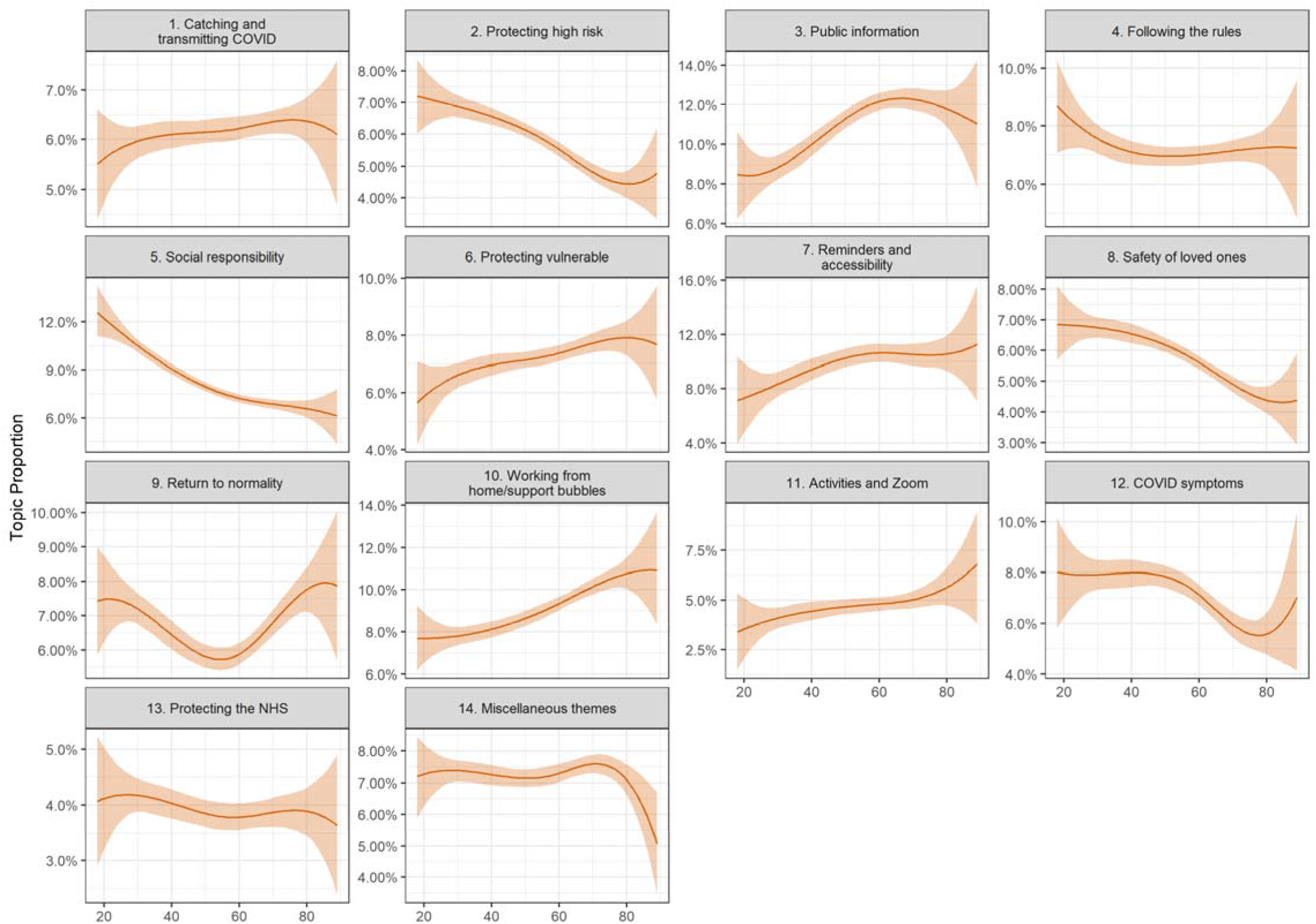
Association between facilitator document topic proportion and participant’s age (+ 95% confidence intervals). Derived from OLS regression models including adjustment for gender, ethnicity, age, education level, living arrangement, psychiatric diagnosis, long-term physical health conditions, self-isolation status, Big-5 personality traits and confidence in government.

**Figure 3:**
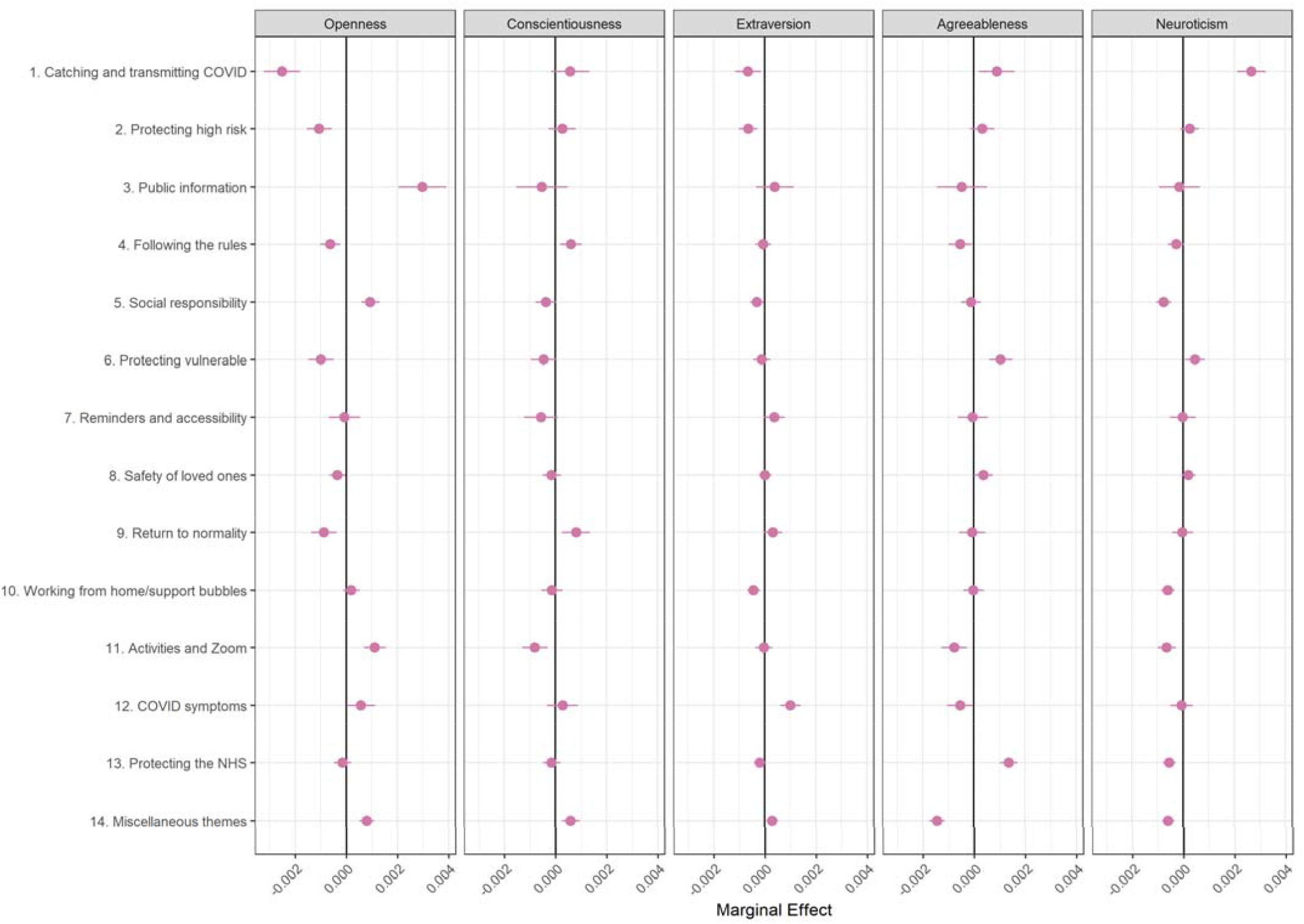
Association between facilitator document topic proportion and Big-5 personality traits (+ 95% confidence intervals). Derived from OLS regression models including adjustment for gender, ethnicity, age, education level, living arrangement, psychiatric diagnosis, long-term physical health conditions, self-isolation status, Big-5 personality traits and confidence in government.

**Figure 4:**
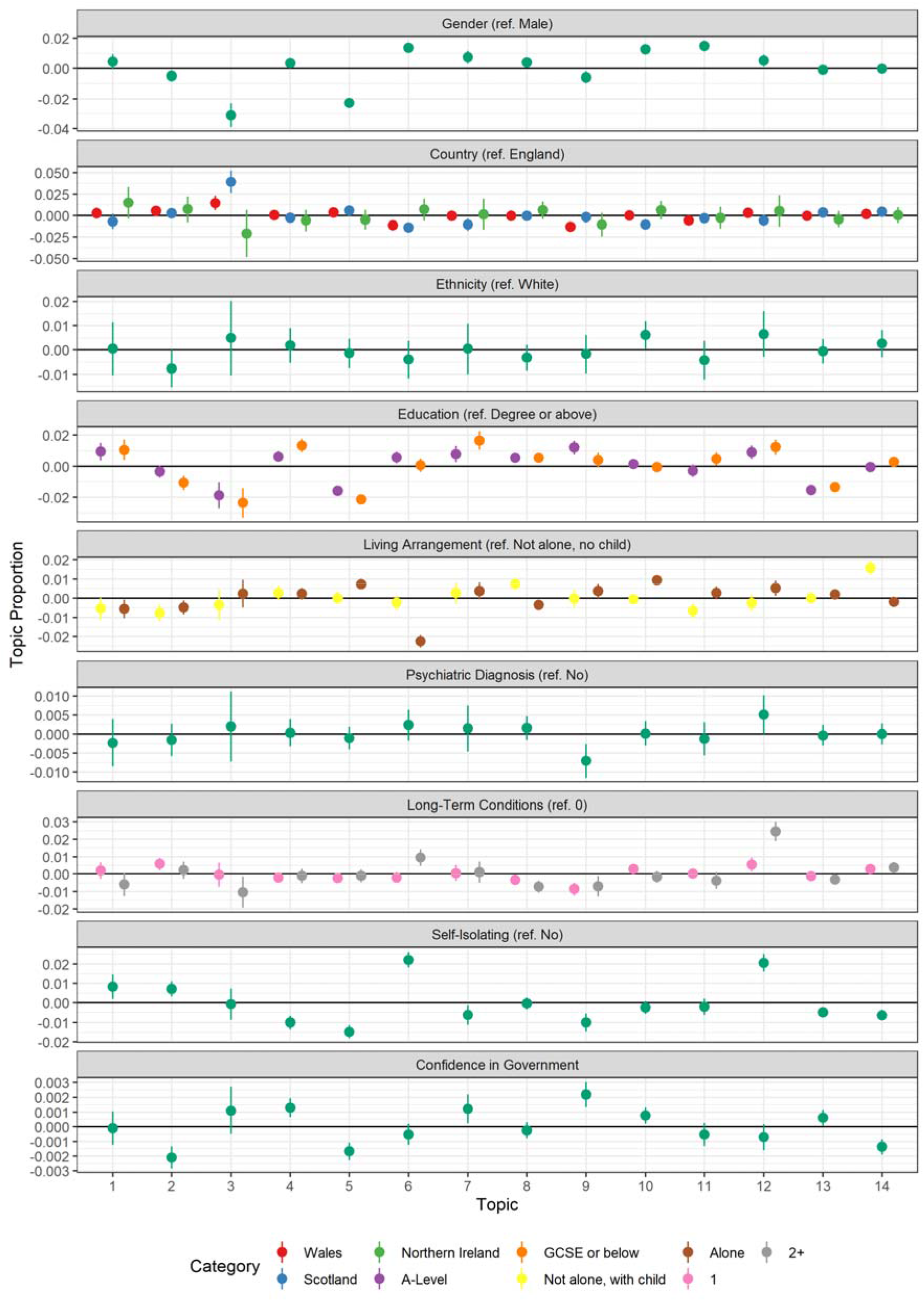
Association between facilitator document topic proportion and participants’ demographic and socioeconomic characteristics, health, and confidence in government (+ 95% confidence intervals). Derived from OLS regression models including adjustment for gender, ethnicity, age, education level, living arrangement, psychiatric diagnosis, long-term physical health conditions, self-isolation status, Big-5 personality traits and confidence in government.

There were a number of differences according to age (Figure 2). Younger participants were more likely to discuss protecting high risk (Topic F2), acting out of social responsibility (Topic F5) and protecting loved ones (Topic F8). Older people were more likely to discuss consuming public information (Topic F3), protecting vulnerable (Topic F6), finding reminders for masks and sanitizer helpful (Topic F7), working from home or using support bubbles (Topic F10), and benefitting from activities and Zoom (Topic F11). Interestingly, there was a U-shaped association between age and Topic F9 (return to normality), with younger and older people more likely to discuss the topic than the middle aged.

There were also several differences according to personality traits (Figure 3). Individuals high in trait openness and extraversion were less likely to discuss catching and transmitting COVID-19 (Topic F1) as a motivation to comply, while more agreeable or neurotic individuals were. Openness was also related to a higher likelihood of discussing public information (Topic F3), acting out of social responsibility (Topic F5), and finding activities and Zoom helpful (Topic F11). Conscientious individuals were more likely to discuss following the rules (Topic F4) or wanting to return to normality (Topic F9), while agreeable individuals were more likely to discuss protecting the vulnerable (Topic F6) and protecting the NHS (Topic F13). Finally, neurotic individuals were less likely to discuss social responsibility (Topic F5), working from home / support bubbles (Topic F10), activities and Zoom (Topic F11), and protecting the NHS (Topic F13), though this may partly be due to the strongly association between neuroticism and discussing catching and transmitting COVID-19 (Topic F1).

Finally, there were several differences according to participants’ demographic and socio-economic characteristics, health and confidence in government (Figure 4). Female participants were less likely to discuss Topic F3 (Public information) or Topic F5 (Social responsibility), but more likely to mention Topic F6 (Protecting vulnerable), Topic F10 (Working from home/support bubbles) or Topic F11 (Activities and Zoom). Individuals with less than degree level education were more likely to discuss Topic F4 (Following the rules) and Topic F7 (Reminders and accessibility) and less likely to discuss Topic F3 (Public information), Topic F5 (Social responsibility), or Topic F13 (protecting the NHS). Individuals who lived alone or who had not been self-isolating were less likely to mention protecting the vulnerable (Topic F6) and individuals with psychiatric diagnoses were more likely to discuss returning to normality as a motivator (Topic F9). Individuals with greater confidence in government were more likely to discuss following the rules (Topic F4) and wanting to return to normality (Topic F9). They were also less likely to mention protecting high risk (Topic F2) or acting out of social responsibility (Topic F5).

### Barriers to Compliance

We selected a 14-topic solution for the responses on barriers to compliance. Short descriptions are displayed in Table 3, along with exemplar quotes and topic titles that we use when plotting results. Three topics related to difficulties complying due to the actions of other people. The largest topic (Topic B1; 20.82%; Others invading space) identified responses on difficulties social distancing in public places (i.e., remaining 2 metres apart) due to others getting too close, particularly in supermarkets or on pavements. Topic B8 (6.43%; Workplace issues) related to issues in workplaces that prevented people from being able to comply. The topic surfaced several responses from teachers describing the challenge of maintaining social distancing with hundreds of schoolchildren. Topic B10 (5.74%; Social norms and social pressures) related to challenges resisting social pressure to break rules from family and friends, and well as the specific demotivation of seeing non-compliance among the general public and members of the government.

**Table 3:** Topic descriptions – barriers to compliance

| Topic | Proportion | Short Title | Description | FREX | Exemplar Text |
| --- | --- | --- | --- | --- | --- |
| B1 | 20.82% | Others invading space | Difficulty of maintaining social distancing in public, due to actions of others, particularly in supermarkets. | space, supermarket, distanc, maintain, social, difficult, other, observ, street, crowd | “Whilst food shopping I keep my distance from others.. however many people do not keep their distance from me.<br>Unbelievable how many people reach over /past me to reach something from shelves i am stood in front of” |
| B2 | 9.79% | Issues with masks and sanitiser | Forgetting to wear masks, wash hands, or use hand sanitiser.<br>Discomfort caused by masks or santiser. | mask, wear, hand, forget, wash, sanitis, deliveri, breath, facemask, dog | “Shortness of breath and asthma means that I don't wear a face covering but I do wear an exemption badge.”<br>“Forgetfulness: workmen renovating my flat nearby, occasionally have forgotten to wear a mask or wash/gel hands - re latter often remember some time after but worry it is too late” |
| B3 | 7.91% | Mental health and family support | Impacts on own and family members mental health and insufficiency of support bubbles to meet practical needs. | mental, health, support, mother, bubbl, elderli, childcar, relationship, dad, carer | “Single parent with twin toddlers, need support from more than one bubble for childcare and to support mental health,”<br>“Mental health issues for myself and relatives. My daughter struggles with not being able to see her boyfriend.” |
| B4 | 7.24% | Special circumstances | Breaking the rules on occasion, usually due to necessity or emotional wellbeing | hous, week, grandchildren, 3, birthdai, grandson, old, 6, hug, 7 | “I look after my grandchildren so kissing and hugging happens, my daughters had their birthdays so we visited to give presents and hugged them, terrible to feel guilty hugging your children. My 85 year old father asks for shopping occasionally, we take it to him and enter the house but feel guilty doing so.”<br>“I look after my grandson 1 day a week & babysit at other times. I have 2 adult sons who moved out recently & come back to the house to get things.” |
| B5 | 7.07% | Following the guidelines | Following the guidelines completely to the best of one’s knowledge and ability. | guidelin, follow, obstacl, transport, line, public, easi, selfish, guid, complet | “I have tried to follow guide lines 100% but have probably slipped up a few times”<br>“I always follow the guidelines.” |
| B6 | 6.56% | Missing family and friends | Desire to meet family and friends. Meeting outside made harder due to poor weather. | friend, miss, garden, famili, christma, allow, meet, spend, coffe, socialis | “Missing family. In summer the weather made it easier and we could meet in the garden, this isn't so tempting in winter!” |
| B7 | 6.55% | Lack of trust in government | Lack of trust and confidence in government competence. Anger at Dominic Cummings. | advic, messag, cum, trust, domin, minist, bori, faith, incompet, johnson | “Boris Johnson, lack of leadership, lack of decisive and timely advice, mixed messages, Dominic Cummings idiocy, continuing lies from Ministers”<br>“lack of confidence in the evidence base for decision-making, lack of confidence that decision-makers are acting in best interests of population” |
| B8 | 6.43% | Workplace issues | Difficulty complying with rules in the workplace, especially in schools. | school, teacher, work, colleagu, primari, student, offic, teach, workplac, meter | “The rules being had to implement in a work environment (secondary school) it's not possible to remain 1metre away from pupils in corridors or in classrooms when there's 30+ in the class”<br>“I am considered an essential worker and have to go to work. Although some of us could work from home we all have to attend the office every day and mix with colleagues.” |
| B9 | 6.39% | Perceiving the risks as low | Belief that the risk is low or that the statistics are inaccurate or exaggerated.<br>Also contains text on difficulties with testing or the Test and Trace system. | covid, test, low, catch, viru,<br>risk, life, symptom, posit,<br>normal | “I am in an area with very low infection rates and still don't think I know anyone who has actually tested positive or been very ill.”<br>“I don't believe the stats in the media and the risks associated with COVID-19 have been completely over exaggerated and are killing the economy. Deaths in hospital are falsely being recorded as COVID-19 and the percentages of people who have died are tiny and do not warrant lockdowns. The few people I know who have tested positive, have not even known they had it. I have zero trust in any facts presented about COVID-19 and therefore I am not going to stop seeing my loved ones.” |
| B10 | 5.74% | Social norms and social pressures | Pressure from family, friends and colleagues to bend rules.<br>Observation of others – including those in power – breaking rules.<br>Also, difficulties related to confusion around the rules. | pressur, stick, break, relax, make, peer, habit, guidanc, rule, tempt | “I have been following the guidelines but I am always made to feel like grinch by colleagues who bend rules to suit themselves. Its (sic) hard”<br>“I have been following guidelines, but I have seen so many people not following the guidelines it makes me more tempted to 'bend the rules' slightly.” |
| B11 | 4.35% | Complexity of rules | Difficulties following frequently changing rules that are also perceived as lacking logical basis. Lack of flexibility in rules. | sens, chang, tier, common, logic, complic, stupid, ridicul, nonsens, appli | “Stupid nonsensical non-evidenced government knee jerk reactions eg closing places of worship and golf clubs with no real rationale.”<br>“I think we should be using common sense rather than overly prescriptive rules that don't always make sense” |
| B12 | 4.16% | Loneliness | Impact on mental wellbeing of living alone. | live, alon, human, interact, limit, loneli, person, contact, period, lowrisk | “I need face to face human contact for my mental well being and my sanity”<br>“I live on my own, and so do the majority of people I interact with. The restriction to meet as two households outside means I can really only ever see one other person, and that really limits meaningful shared experiences which I think are important for my mental health. I have on occasion broken this rule to meet e.g. as three of us outside.” |
| B13 | 3.54% | Geographical variation in rules | Difficulties related to differences in rules across areas, particularly for those living on the border between England and Wales. | england, wale, border, understand, letter, attitud, action, measur, interpret, consist | “I live right on the border between Wales and England and have not been able to stay entirely within Wales at times when the border was closed because of the locations of essential local services”<br>“Frequency of changes to guidelines. Differences between devolved and English measures. English measures have been more clearly communicated. I'm still uncertain on Welsh rules for meeting other people.” |
| B14 | 3.45% | Confusion around rules | Confusion about what the rules are.<br>Also contains text on adhering with guidelines to best of one's ability. | hinder, frustrat, applic, regul, awar, adher, situat, confus, abil, disregard | "The instructions are confused and confusing. The variety of scientific opinion is not always reflected but interviewers seem sometimes more interested in apportioning blame than establishing consensus." |

Several topics related to difficulty understanding current rules. Topic B11 (4.35%; Complexity of rules) identified individuals who had difficulty keeping abreast of (frequently changing) rules or who could not understand the logic behind the rules (for instance, keeping pubs open but stipulating only four people could meet). Some responses identified in this topic also questioned the lack of flexibility in rules or the allowance for individuals to apply common sense. Topic B13 (3.54%; Geographical variation in rules) related to challenges arising from the variation in rules around the UK, particularly for those living near borders, such as that between England and Wales. One frequently noted issue was accessing essential services located on the other side of a border. Topic B14 (3.45%; Confusion around rules) included responses that described general confusion about what the rules were, though this topic also identified individuals who stated following guidelines to the best of their ability. (Topic B5 [7.07%; Following the guidelines] also related to individuals stating that they had been following guidelines completely.) Criticism of the government was voiced in Topic B7 (6.55%; Lack of trust in government), with individuals noting a lack of confidence or trust in the government’s decisions or motives, including in Boris Johnson’s leadership. Several participants also expressed anger at Dominic Cummings – a senior Government advisor – and the decision to keep him in post following reports in May 2020 that he had broken lockdown rules.

Several topics related to the negative impact of compliance on the lives of participants and their family and friends. Topic B3 (7.91%; Mental health and family support) related directly to the impact of restrictions on mental health for the participant themselves and their family and friends. The topic also identified responses from individuals who had extended support bubbles beyond the rules to provide or receive support to loved ones. This included both emotional and practical support (e.g., by providing childcare). Topic B6 (6.56%; Missing family and friends) related to difficulties due to missing family and friends, particularly when the weather turned poor and meeting outside became a challenge. Topic B12 (4.16%; Loneliness) identified responses from individuals who lived alone and had struggled due to the resulting social isolation. Some individuals described breaking lockdown rules on occasion for human contact. Topic B4 (7.24%; Special circumstances) also identified individuals who had occasionally broken lockdown rules to improve wellbeing, but also identified individuals breaking rules to provide support to family members (e.g., childcare) or out of temporary necessity (e.g., helping a family member move out of their home).

Practical barriers to compliance were reflected in Topic B2 (9.79%; Issues with masks and sanitiser) which related to difficulties with face masks or using hand sanitiser. Several respondents stated forgetting to wear masks or use sanitiser or noted discomfort from the use of these due to existing health conditions. Finally, Topic B9 (6.39%; Perceiving the risks as low) identified individuals who did not comply with guidelines due to perceptions that risks were low, for instance, due to previous infection with COVID, low caseloads in the local area or beliefs that government statistics were exaggerated. The topic also identified a number of responses of individuals who had not gotten tested when displaying symptoms, either due to lack of availability or beliefs that PCR tests were not accurate.

### Compliance Barriers and Respondent Characteristics

Figures 5-7 display the results of models regressing topic proportions on respondent characteristics. There were a number of differences according to age (Figure 5). Older participants were more likely to discuss issues with masks and sanitiser (Topic B2) and missing family and friends (Topic B6). They were also less likely to discuss special circumstances acting as barriers to following rules (Topic B4), having a lack of trust in government (Topic B7), perceiving COVID-19 to be of low risk (Topic B9), or describing issues with the geographical variation in rules (Topic B13). Young people were more likely to mention social pressures (Topic B10) and middle-aged people were least likely to discuss loneliness and social isolation (Topic B12).

**Figure 5:**
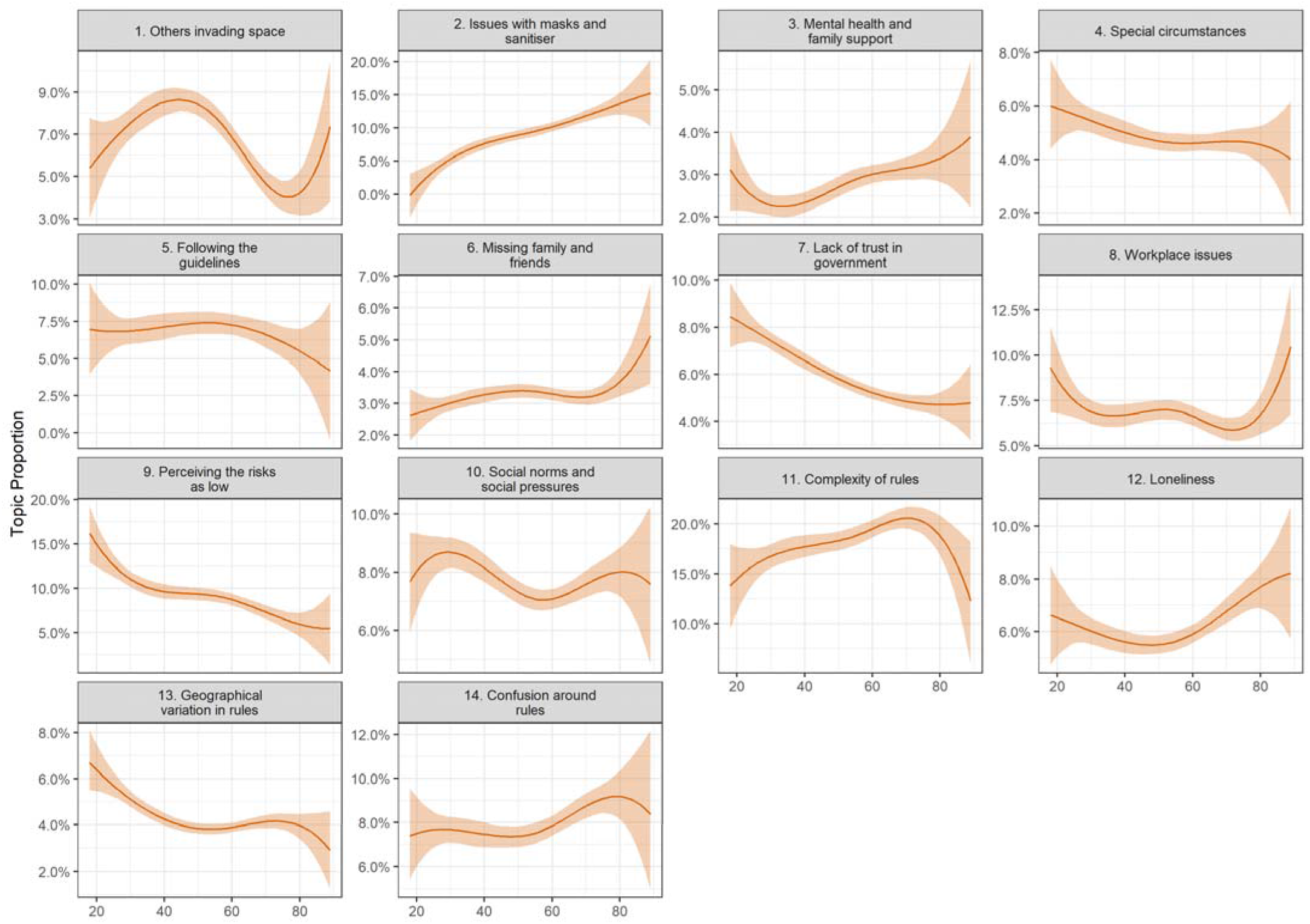
Association between barrier document topic proportion and participant’s age (+ 95% confidence intervals). Derived from OLS regression models including adjustment for gender, ethnicity, age, education level, living arrangement, psychiatric diagnosis, long-term physical health conditions, self-isolation status, Big-5 personality traits and confidence in government.

There were also a number of differences according to personality traits (Figure 6). Notably, extraverted individuals were less likely to discuss others invading space (Topic B1), having issues with masks or sanitizer (Topic B2) or following the guidelines completely (Topic B5) but were more likely to mention social factors such as mental health and family support (Topic B3), breaking guidelines on occasion (Topic B4), missing family and friends (Topic B6), difficulties with social norms and social pressures (Topic B10), and loneliness (Topic B12). Neurotic individuals were less likely to discuss COVID as being low risk, and conscientious individuals were more likely to discuss following the guidelines completely (Topic B5).

**Figure 6:**
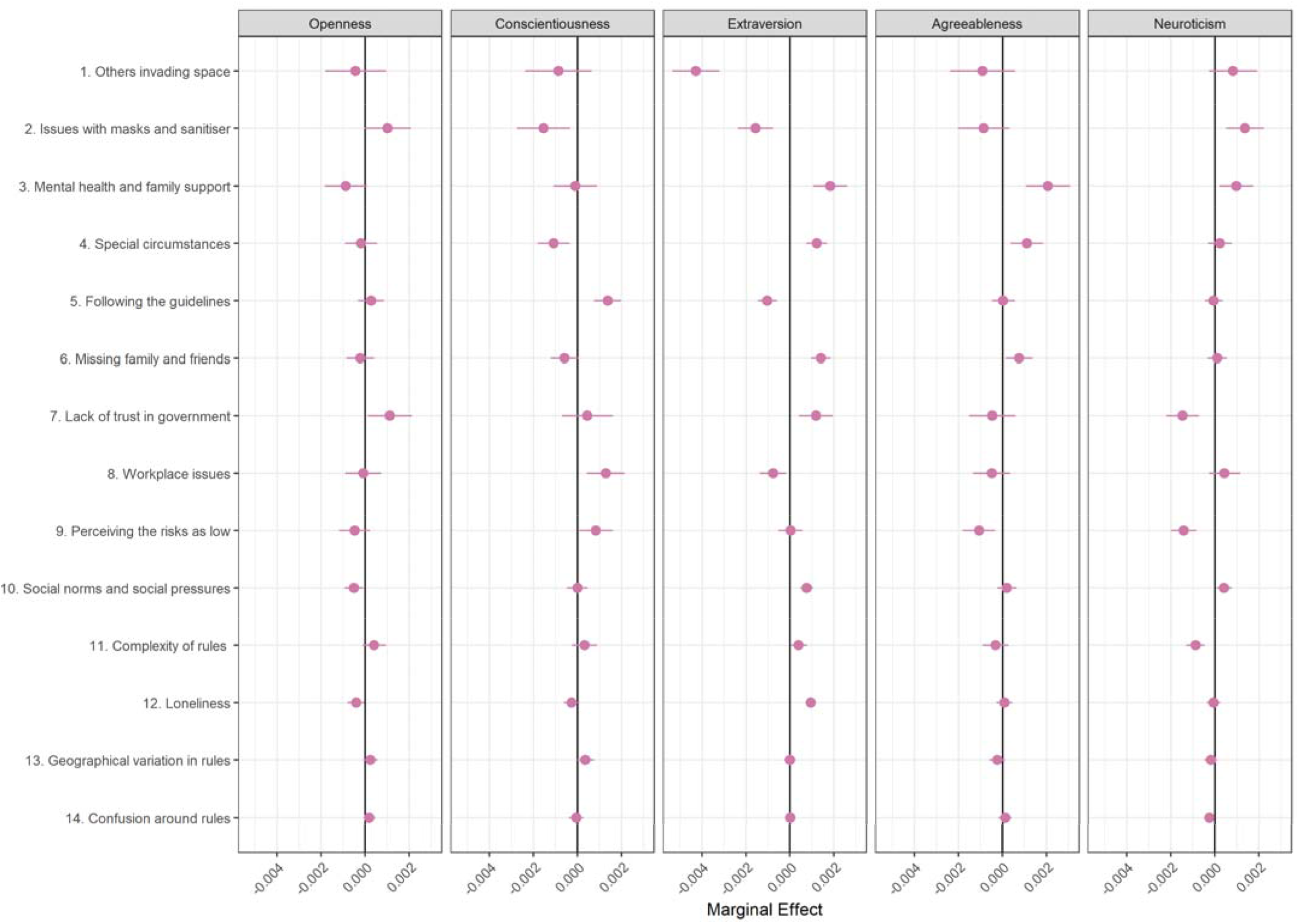
Association between barrier document topic proportion and Big-5 personality traits (+ 95% confidence intervals). Derived from OLS regression models including adjustment for gender, ethnicity, age, education level, living arrangement, psychiatric diagnosis, long-term physical health conditions, self-isolation status, Big-5 personality traits and confidence in government.

Finally, there were several differences according to participants’ demographic and socio-economic characteristics, health and confidence in government (Figure 7). Female participants were more likely to discuss Topic B3 (Mental health and family support), Topic B4 (Special circumstances), and Topic B5 (Following the guidelines) and less likely to discuss lack of trust in government (Topic B7). Individuals with less than degree level education were also more likely to state they were following the guidelines completely (Topic B5) but were less likely to discuss missing family and friends (Topic B6). Individuals with children were more likely to discuss workplace issues (Topic B8), while individuals with psychiatric diagnoses were more likely to mention others invading space (Topic B1) or having issues with masks and sanitizer (Topic B2). Supporting our structural topic models, individuals with high confidence in government were less likely to discuss Topic B7 (lack of trust in government) and individuals who lived alone were more likely to discuss loneliness (Topic B12).

**Figure 7:**
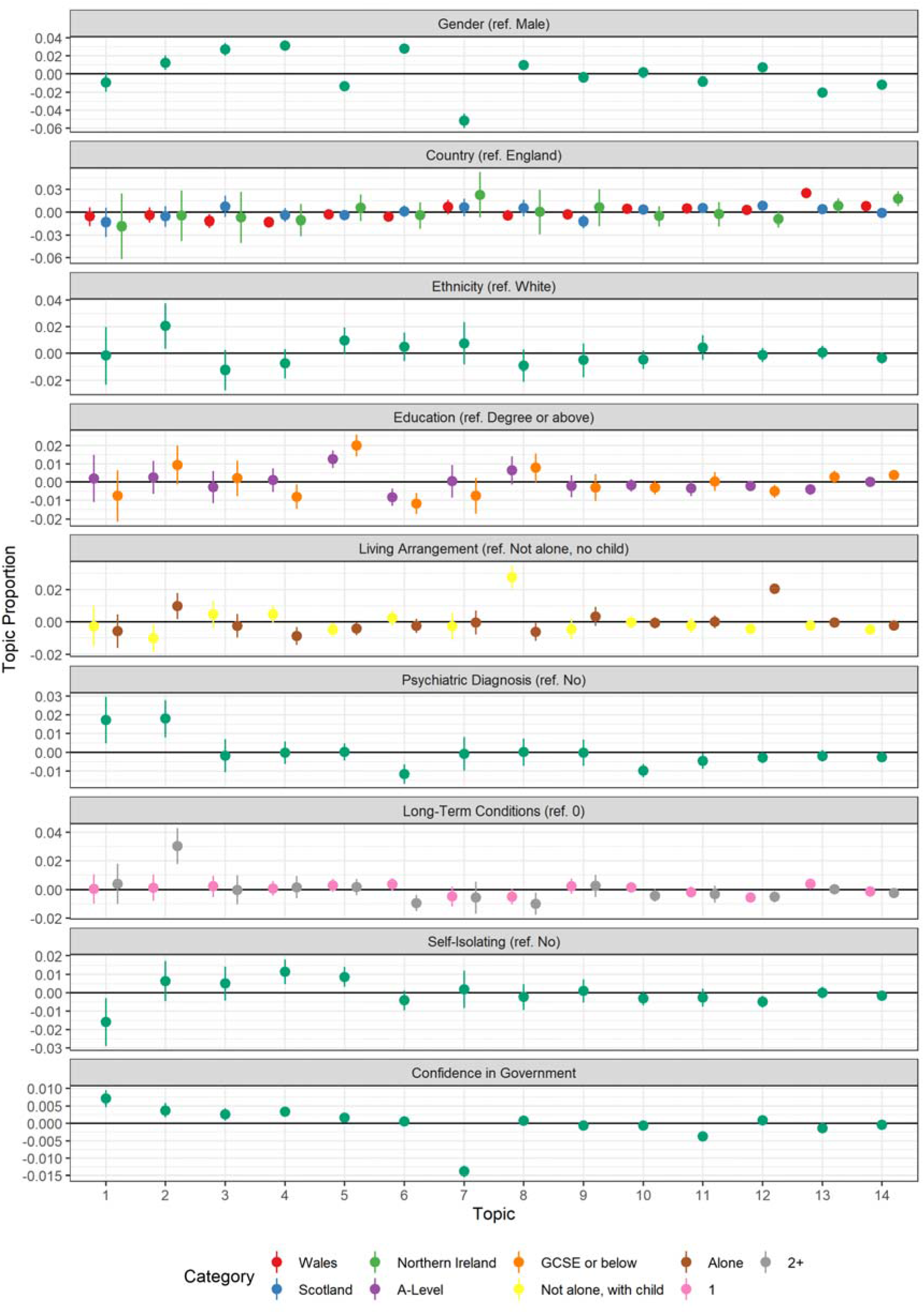
Association between barrier document topic proportion and participants’ demographic and socioeconomic characteristics, health, and confidence in government (+ 95% confidence intervals). Derived from OLS regression models including adjustment for gender, ethnicity, age, education level, living arrangement, psychiatric diagnosis, long-term physical health conditions, self-isolation status, Big-5 personality traits and confidence in government.

### Associations Between Topic Proportions and Self-Reported Compliance

The results of regressions estimating the average level of self-reported compliance according to topic proportions are displayed in Figure 8. (The dashed line represents the mean compliance level across the relevant sample.) The top panel shows results for facilitators, the bottom for barriers to compliance.

**Figure 8:**
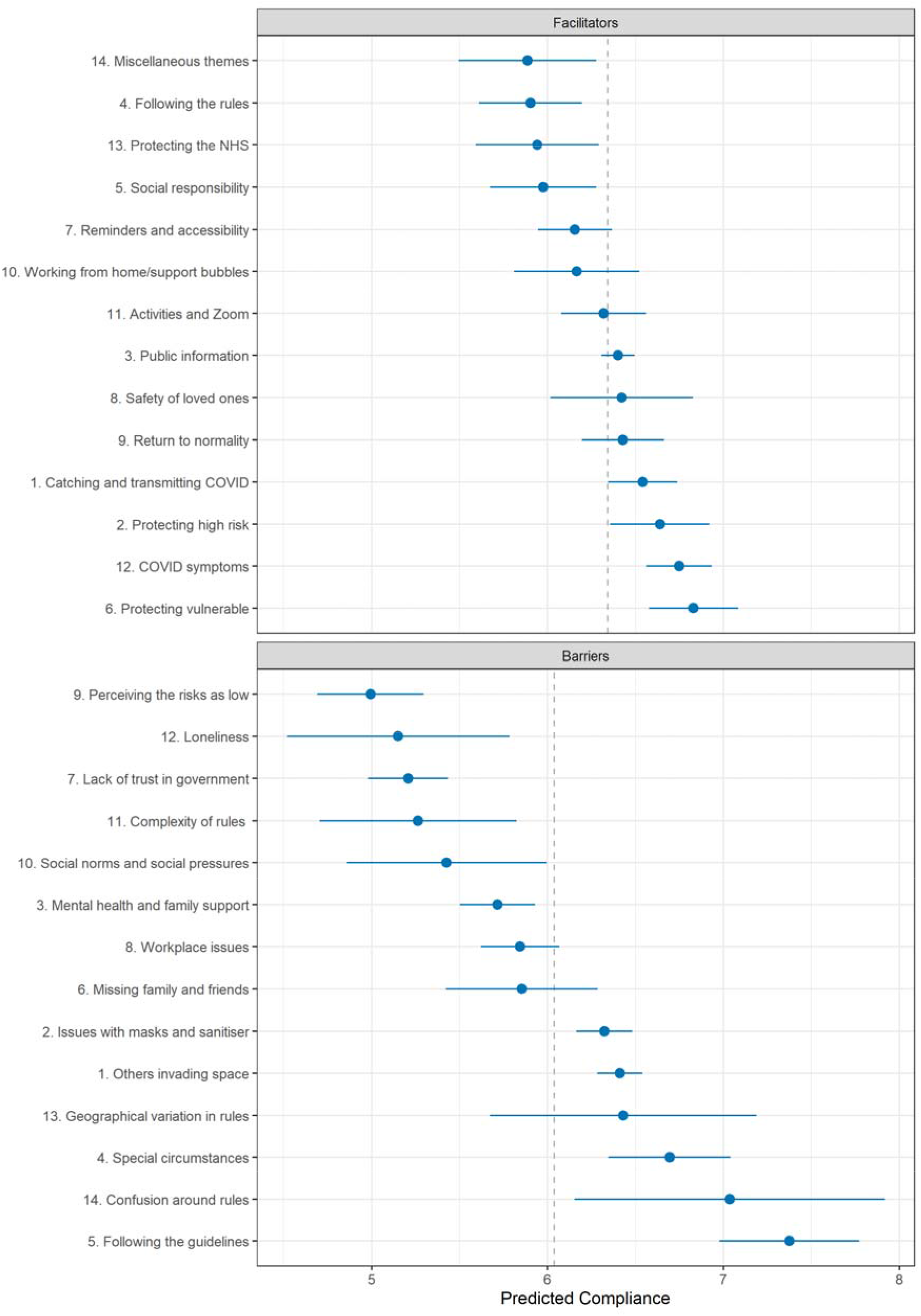
Association between self-reported compliance with COVID-19 related guidelines and document topic proportions (+ 95% confidence intervals). Dashed line indicates means compliance levels in sample used in regression.

Facilitator topics related to desires to protect the vulnerable (Topic F6) or high risk (Topic F2) or reduce the likelihood of catching and transmitting COVID-19 were associated with highest compliance levels. Following the rules (Topic F4), desire to protect the NHS (Topic F13) and acting out of social responsibility (Topic F5) were related to lowest average compliance levels. For barriers to compliance, perceiving COVID-19 as representing a low risk (Topic B9), living alone (Topic B12), lack of trust in government (Topic B7) and finding the rules to be too complex (Topic B11) was related to lowest compliance levels. Following the guidelines completely (Topic B5), confusion around the rules (Topic B14) and facing special circumstances (Topic B4) were related to higher than average compliance.

## Discussion

Using free-text data from 17,000 adults, we identified several facilitators and barriers to compliance with COVID-19 guidelines, eight months after lockdown measures were implemented in the UK. For facilitators of compliance, a sizeable proportion of text was related to desire to reduce risks for oneself, one’s family and friends, and – to a lesser extent – the general public and the NHS and its workers, specifically. Some participants also spoke of being motivated by a sense of responsibility, a desire to return to life as normal, or acting from a predisposition to follow rules in general. For barriers to compliance, a substantial proportion of text was related to other people making compliance difficult, either by getting too close in public or workplaces, putting social pressure on participants to violate guidelines, or acting as a demotivator when their non-compliance was observed. Participants also spoke of the emotional toll of complying with guidelines, particularly among those who lived alone, missing family and friends, and of the necessity of providing support to loved ones. Interestingly, participants also spoke of breaking rules on occasion, for instance when social isolation had gotten too great. A large portion of text was also devoted to issues with the guidelines themselves. Participants found guidelines confusing and often did not see their logical basis. The variation in rules across time and geographic areas was a particular issue. Some participants also discussed a lack of trust in government and expressed anger at the decision to keep Dominic Cummings in his position as government advisor after he broke lockdown rules. Last, some participants discussed believing COVID-19 to be low risk as a reason for not complying. Individuals who spoke about this had the lowest self-reported compliance overall.

In Table 4, we map the topics identified in this analysis to components of the COM-B framework [23, 24]. Most of the enablers to comply were related to reflective motivation that complying would reduce adverse events and support in the return to normality. People also reported that compliance was enabled when there were clear physical opportunities to comply. Barriers to compliance were also related to reflective motivations but also to psychological capabilities and lack of social opportunity. This suggests that behaviour change techniques such as education (e.g. increasing understanding about the virus), persuasion (e.g. stimulating action through inducing positive emotions around the benefits of compliance to society), and incentivisation (e.g. communicating how better compliance could lead to lower virus levels and less need for strict measures) could help to improve motivation, whilst clearer rules, identification and removal of factors hindering compliance, and restructuring of environments (such as shops) to facilitate compliance could help to address the barriers identified [23].

**Table 4:**
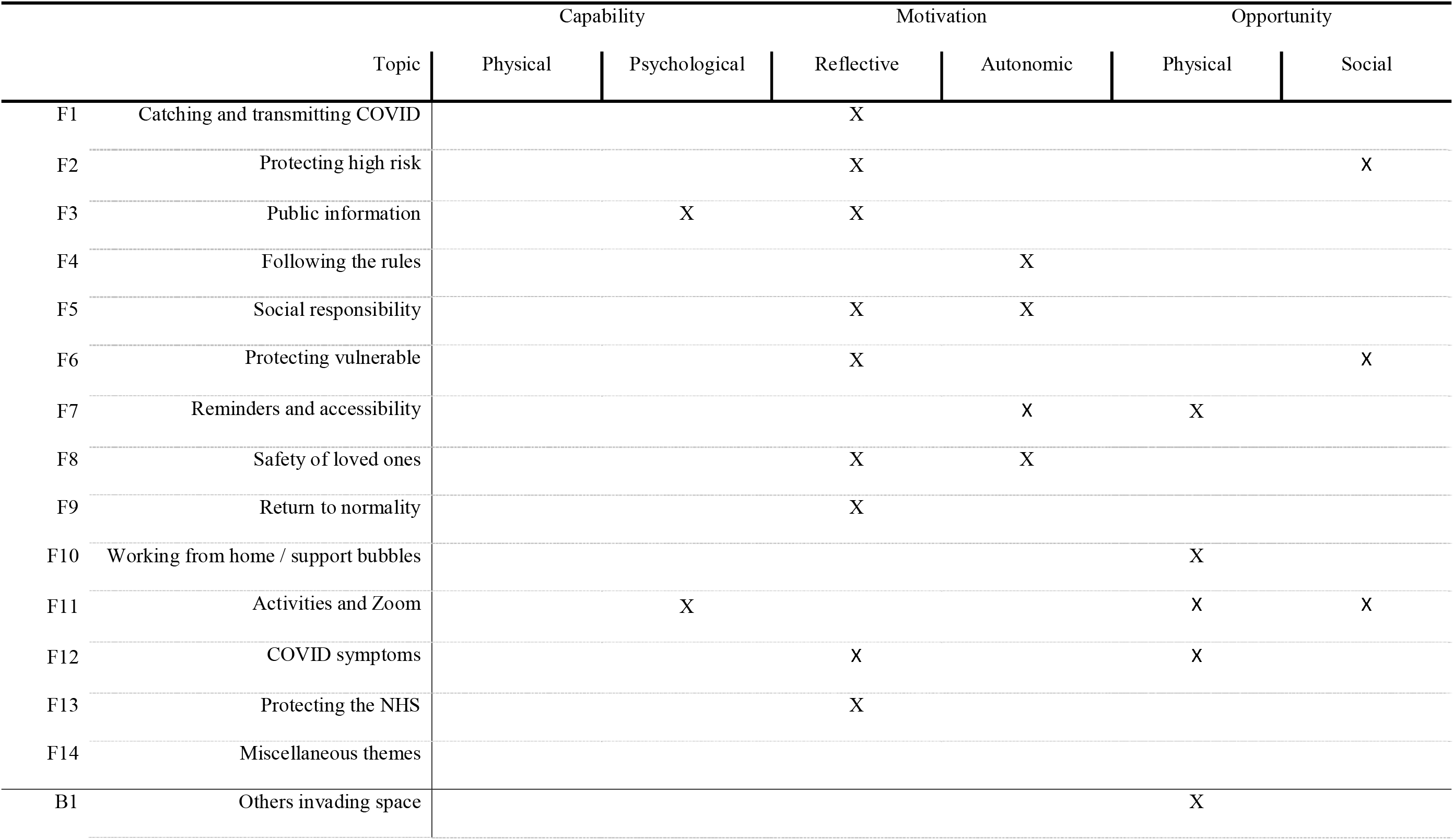
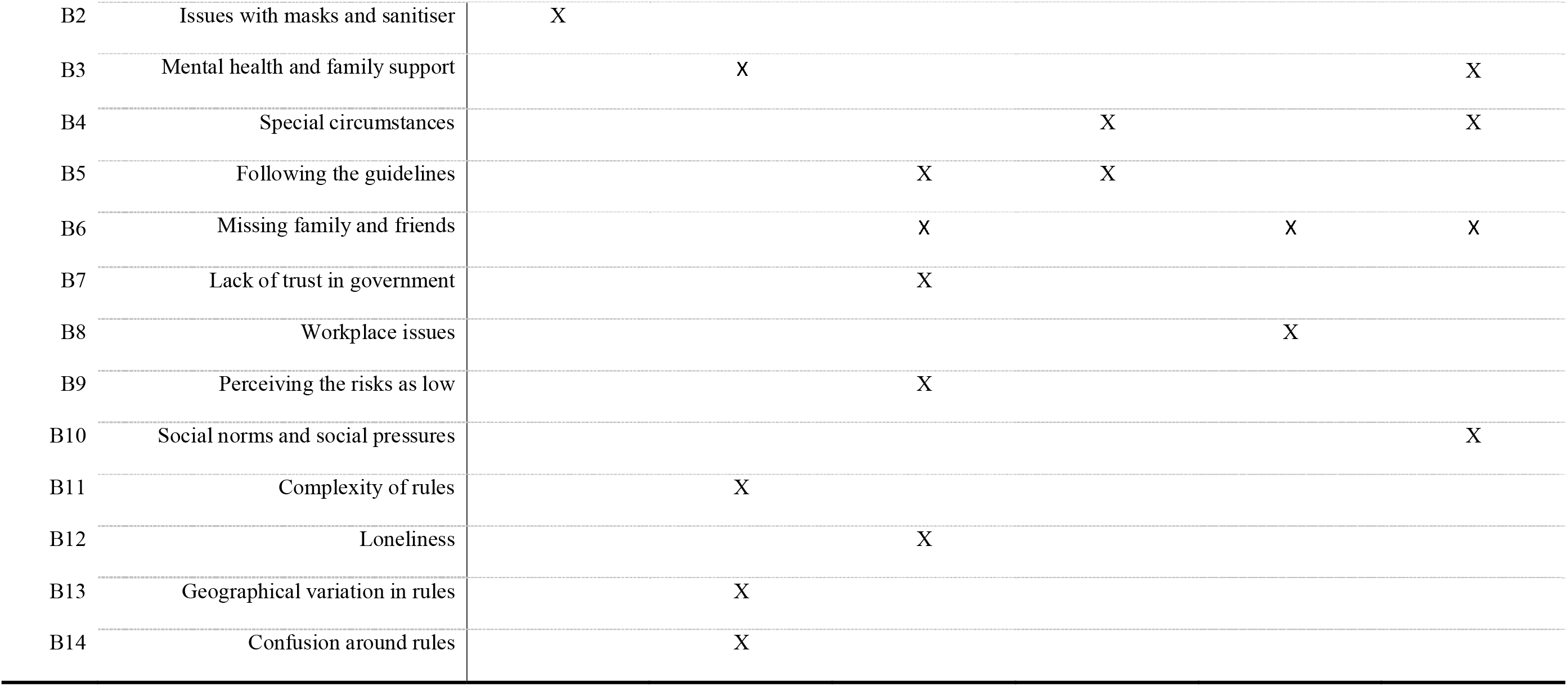
Mapping of topics onto COM-B framework

The topics that participants discussed were related to participant characteristics. Notably, middle aged individuals were least likely to discuss complying out of a desire to end the pandemic and return to normalcy, and younger individuals were more likely to mention acting out of a sense of social responsibility. Extraverted individuals and women were more likely to report emotional challenges and lack of social contact as a barrier to compliance, while conscientious individuals were more likely to state complying with guidelines completely.

Our results are consistent with those of Coroiu et al. [16], who study the barriers and facilitators of compliance with COVID-19 social distancing and shelter-in-place rules. The authors find that a desire to protect oneself and others and acting out of a sense of social responsibility are among the most endorsed items on motivations to comply. They also find that, among the barriers to compliance, believing the risk of catching COVID-19 to be small, not trusting government messaging, need to provide support to friends or family, and feeling stressed when socially isolation are the most endorsed factors. We add to their results by finding that protecting family or friends appears to be a more important motivation than protecting wider society and that confusion about specific rules is a major barrier to compliance.

Our finding that a number of participants violated guidelines on occasion for emotional reasons is important in light of debates on behavioural fatigue [4, 9, 10, 12]. Existing tests of behavioural fatigue have assumed that fatigue would lead to decreasing motivation to comply across the pandemic [4, 5], a test that is in line with what England’s Chief Medical Officer, Chris Whitty, seemingly had in mind when discussing the concept (“Once we have started these things we have to continue them through the peak, and there is a risk that, if we go too early, people will understandably get fatigued and it will be difficult to sustain this over time.” [8]). Our results indicate that some individuals violate rules after a period of sustained compliance, but return to complying, a process akin to resting after a workout [9]. The implication of this may be that individuals can comply over extended periods, if occasional opportunities to “reset” are available.

Our finding that many participants had struggled to understand or keep abreast of changing rules (“alert fatigue”) was consistent with previous qualitative work showing difficulties understanding government messaging [28, 29, 37]. The results suggest that simplified rules may improve compliance, though this would have to be balanced against the cost of keeping some individuals under strict measures for longer than necessary. The lack of a clear rationale for certain rules was cited as reducing motivation to comply suggests that the UK government could increase the transparency of its decision making and that this may improve the ability to tackle COVID-19. Our finding that low confidence in government was a barrier to compliance was consistent with several previous quantitative studies [25–27] and is concerning in light of the decrease in confidence in the UK government that occurred from the beginning of the pandemic to the time of the study [5, 38], but also offers hope for future compliance given the increase in confidence following the rollout of the vaccine [38].

Also of policy interest was the role of mental health and the need to receive or provide support to family and friends as a barrier to compliance. Social bubbles were not introduced immediately by the governments of UK. The results here suggest this rule may have caused stress or been ignored by some households. Further, some participants complained that support bubbles were of insufficient size when introduced. More flexibility may have improved the wellbeing of individuals requiring support, though an issue is that this flexibility could increase the complexity of rules, which as discussed, can raise its own issues for facilitating high compliance.

This study had several strengths. We used rich qualitative data from over 17,000 UK adults representing a wide range of demographic groups. Using structural topic modelling, we were able to combine qualitative and quantitative approaches, to identify unique facilitators and barriers to compliance with guidelines in the UK, and to assess how response topics differed according to participant characteristics. Our results have several policy implications and showed (to our knowledge) unique evidence of occasional, isolated rule violations among some of the general public. The results also add detail to previous results quantitative results showing a link between age, personality traits and compliance behaviour [5, 14, 21]. Some of the topics were related in the expected direct to participant characteristics – for instance, individuals with low confidence in government were more likely to offer criticisms of the government – which suggests that the structural topic models extracted consistent and meaningful themes. Further, we used data from 8-9 months after the first lockdown, later than much of the literature on compliance during COVID-19, which has focused on the early stages of the pandemic.

Nevertheless, this study had several limitations. Not all of the topics identified a single theme consistently. Associations with participant characteristics could be driven or biased by idiosyncratic texts. We used a convenience sample that, though heterogeneous, was unrepresentative of the UK population and, further, respondents to the free-text question were biased towards the highly educated. Response biases could also have generated bias in the topic regression results. As we focused on topics offered spontaneously by respondents, a participant not writing about a particular topic does not necessarily imply that the participant does not agree with its sentiment, though our assumptions is that the proportion of text devoted to a topic is related to its perceived importance. Finally, while we included a wide set of predictors in our structural topic models, many relevant factors were unobserved. Associations may be biased by unobserved confounding.

## Conclusions

We identified several facilitators and barriers to compliance. Of particular importance for facilitating compliance was concerns about the risk of COVID-19 for one’s self and one’s family and friends, while important barriers to compliance were problems maintaining social distance in public spaces and difficulties understanding and keeping abreast of government rules. The results suggest that government communications that emphasizes the potential risks of COVID-19 and provides simple, consistent guidance on how to reduce the spread of the virus would improve compliance with preventive behaviours. Investments in managing or reorganizing public space – particularly in supermarkets – so that social distancing is encouraged could also have significant positive effects. While a large literature has related individual characteristics to preventive behaviour, our results give fresh insight into the wider contextual issues that are important for compliance.

## Data Availability

Due to stipulations set out by the ethics committee, the free-text data cannot be made publicly available. The code used in the analysis is available at https://osf.io/nf4m9/.

## Declarations

### Ethics approval and consent to participate

The authors assert that all procedures contributing to this work comply with the ethical standards of the relevant national and institutional committees on human experimentation and with the Helsinki Declaration of 1975, as revised in 2008. The study was approved by the UCL Research Ethics Committee [12467/005] and all participants gave informed consent.

### Consent for publication

Not applicable.

### Availability of data and materials

The free-text data used in this study cannot be made publicly available due to stipulations set out by the ethics committee. The code used to run the analysis is available at https://osf.io/nf4m9/.

### Competing interests

The authors declare that they have no competing interests.

### Funding

This Covid-19 Social Study was funded by the Nuffield Foundation [WEL/FR-000022583], but the views expressed are those of the authors and not necessarily the Foundation. The study was also supported by the MARCH Mental Health Network funded by the Cross-Disciplinary Mental Health Network Plus initiative supported by UK Research and Innovation [ES/S002588/1], and by the Wellcome Trust [221400/Z/20/Z]. DF was funded by the Wellcome Trust [205407/Z/16/Z]. The study was also supported by HealthWise Wales, the Health and Car Research Wales initiative, which is led by Cardiff University in collaboration with SAIL, Swansea University. The funders had no final role in the study design; in the collection, analysis and interpretation of data; in the writing of the report; or in the decision to submit the paper for publication. All researchers listed as authors are independent from the funders and all final decisions about the research were taken by the investigators and were unrestricted.

### Authors’ contributions

Data cleaning and analysis was carried out by LW. LW selected the number of model topics and LW, EP, and AS agreed upon narrative titles for the topics. Due to stipulations set out by the ethics committee, data will be made available at the end of the pandemic.

## Acknowledgements

The researchers are grateful for the support of a number of organisations with their recruitment efforts including: the UKRI Mental Health Networks, Find Out Now, UCL BioResource, HealthWise Wales, SEO Works, FieldworkHub, and Optimal Workshop.

## Notes

### Competing Interest Statement

The authors have declared no competing interest.

### Author Declarations

The study was approved by the UCL Research Ethics Committee [12467/005] and all participants gave informed consent.

