## Supplementary Information for "Facilitators and Barriers to Compliance with COVID-19 Guidelines: A Structural Topic Modelling Analysis of Free-Text Data from 17,500 UK Adults"

### Measures

#### Predictors of Topic Proportions

We included the demographic variables for country of residence (England, Scotland, Wales, Northern Ireland), sex (male, female), ethnicity (White, Non-White) and age (basis splines, degrees of freedom 4). We also included three variables for socio-economic position (SEP): education level (GCSE or below, A-levels or equivalent, degree or above) and living arrangement (not alone, no child; not alone, with child; alone). Each variable was measured at baseline interview.

Personality was measured at baseline interview using the Big Five Inventory (BFI-2; Soto & John, 2017), which measures five domains and 15 facets: openness (intellectual curiosity, aesthetic sensitivity, and creative imagination), conscientiousness (organisation, productiveness, and responsibility), extraversion (sociability, assertiveness, and energy level), agreeableness (compassion, respectfulness, and trust) and neuroticism (anxiety, depression, and emotional volatility). Each item was scored on a 5-point scale (1 = “strongly disagree”, 5 = “strongly agree”). We use the sum Likert score for each domain (range 3 - 15). High values indicate high levels of the trait.

Confidence in government was measured with two items: “How much confidence do you have in the CENTRAL UK GOVERNMENT that they can handle Covid-19 well?”; “If you live in a DEVOLVED NATION (i.e. Scotland, Wales or NI), how much confidence do you have in the government WITHIN YOUR OWN COUNTRY that they can handle Covid-19 well?”. Each was measured on a 1 (None at at all) to 7 (Lots) scale. Responsibility for managing the pandemic is devolved to the home nation governments of Wales, Scotland, and Northern Ireland, while the central UK government has powers is responsible for managing the pandemic in England. Thus, we used responses to the first question for participants residing in England and the second question for participants residing in Wales, Scotland, and Northern Ireland. We drew responses from the same data collection as the free-text data.

Finally, we included three variables for participants’ health: pre-existing psychiatric diagnosis, shielding due to (own) pre-existing health conditions, and number of long-term physical health conditions (categorical: 0, 1, 2+). Long-term physical health conditions (0, 1, 2+) was measured using a multiple-choice question on medical conditions. Included conditions were high blood pressure, diabetes, heart disease, lung disease, cancer, any other clinically-diagnosed chronic physical health conditions, or any disability. Psychiatric diagnosis (yes, no) was with the same multiple choice question using items on clinically diagnosed depression, clinically diagnosed anxiety, and any other clinically diagnosed mental health problem. The data was collected at baseline interview. Shielding was defined as not leaving home due to being high risk. Survey items on shielding were included in all data collections from 23 March – 04 July 2020. We defined a person as shielding if they stated shielding at any data collection between these dates.

#### Self-Reported Compliance with Guidelines

Self-reported compliance with COVID-19 guidelines was measured with a single item: “Are you following the recommendations from authorities to prevent spread of Covid-19?”. Responses were scored on a 1 (“Not at all”) to 7 (“Very much so”) scale and were drawn from the same data collection as the free-text data.

### Policy Environment

The period 17 November to 23 December 2020 overlaps with the second wave of COVID-19 in the UK in which there were several changes to COVID-19 related rules. The rules differed somewhat between the nations of England, Scotland, Wales, and Northern Ireland, though mask wearing in indoor public spaces was legally mandated in each nation. On 17 November, England was two weeks into a four-week national lockdown in which non-essential business were closed, employees were asked to work from home where possible, and household mixing was banned. A new three-tier system was put in place from 2 December when lockdown ended. Several areas were placed in the highest tier. In Wales, rules were more relaxed, following a three week “circuit breaker” lockdown that ended on 9 November. Two households were allowed to form bubbles, non-essential shops were open and groups of four people were allowed to meet in indoor public spaces. In Scotland, a five-tier localised system was in place. On 20 November, eleven council areas were placed in tier 4 and non-essential travel between Scotland and England was banned. Northern Ireland was in a circuit breaker lockdown, due to expire on 27 November. Following this, strict rules are put in place, with non-essential shop barred from opening and limited household mixing allowed.

On 24 November, the leads of the four national Governments announced plans to allow mixing of three households for five days between 23-27 December (22-28 December in Northern Ireland), though these plans were later reduced and restricted in some areas. On 2 December, the UK approved the Pfizer/BioNTech COVID-19 vaccine with the first vaccination provided on 8 December. 137,987 people were vaccinated in the first week. On 14 December, a new, more virulent variant of SARS-CoV-2 is identified. Areas in the South East of England are placed into lockdown from 19 December, with Christmas bubbles cancelled in these areas and relaxation elsewhere in England limited to Christmas day. Figure S2 shows 7-day COVID-19 caseloads and confirmed deaths, along with the Oxford Policy Tracker, a numerical summary of policy stringency (Hale et al., 2020), across the study period.

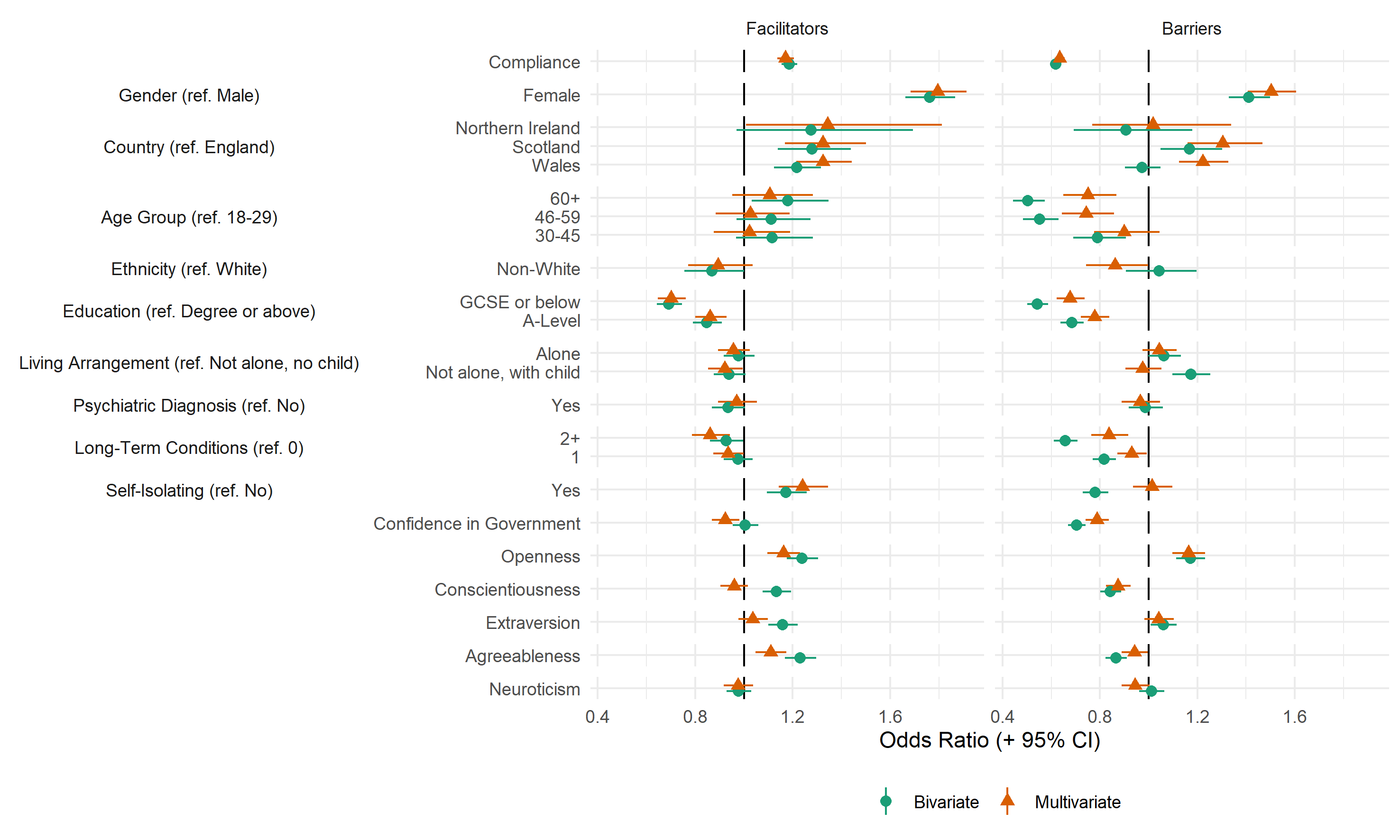

Figure S1: Association between providing a valid free-text response and study variables, derived from bivariate and multivariate logistic regression models.

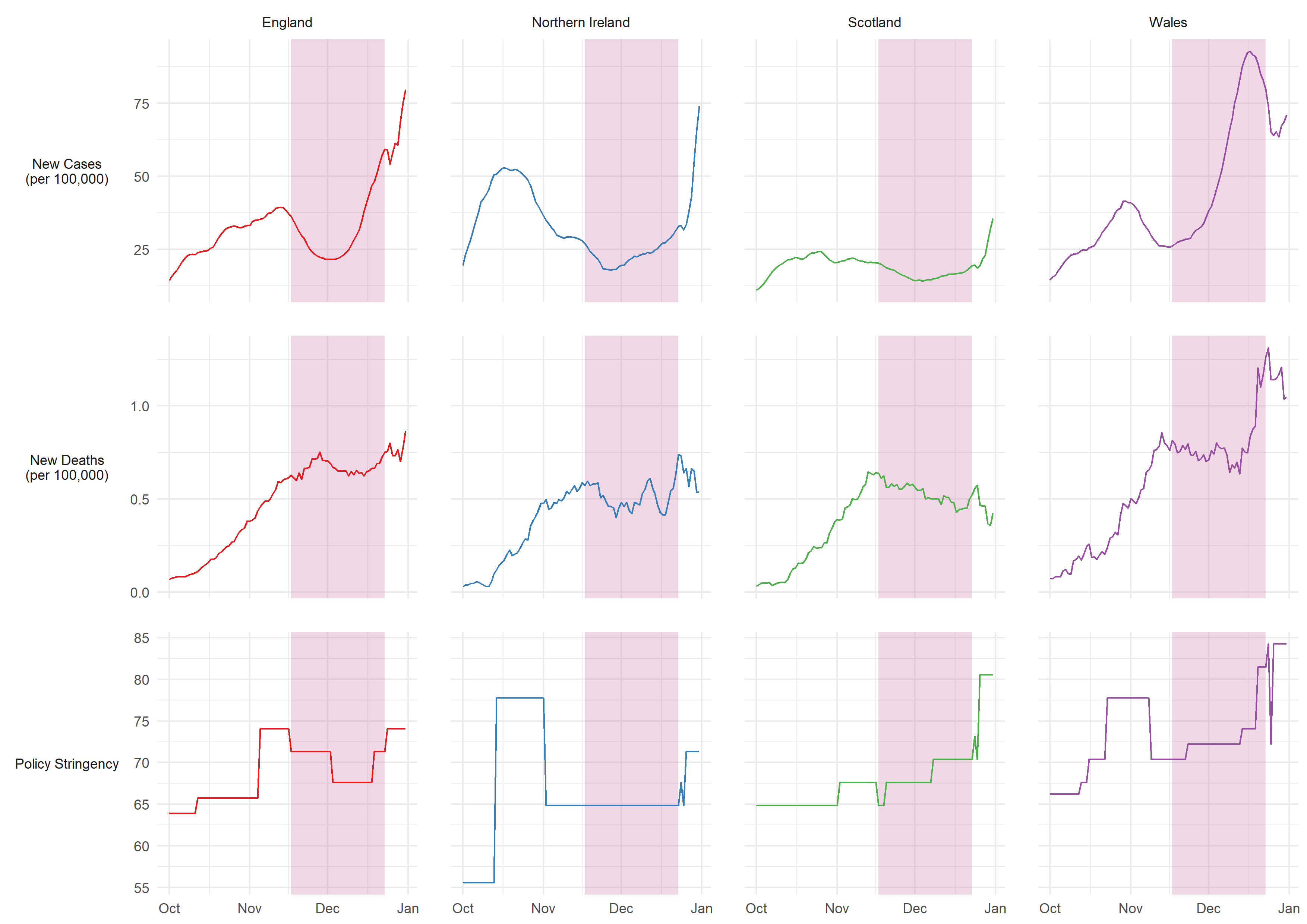

Figure S2: 7-day COVID-19 confirmed cases, COVID-19 deaths, and policy stringency by country. Source: Hale et al. (2020). Pink shaded band represents period in which compliance behaviours were measured.
